# MeshScope-Scenario: A Seeded Monte Carlo Framework for Probabilistic Assessment of ICU and HCU Capacity Shortfall in Japan’s Secondary Medical Areas, Incorporating Inter-Zone Transfer and Seasonal Surge

**DOI:** 10.64898/2026.08.05.26359832

**Authors:** Kunihisa Ohno, Masato Hirai, Satoru Hashimoto

## Abstract

**Background:** Descriptive mapping of intensive care unit (ICU) and high care unit (HCU) capacity across Japan’s secondary medical areas (SMAs) characterizes where beds exist, but medical planning also requires answers to prospective questions: how likely is a capacity shortfall under demand surge, which assumptions drive that risk, and how much protection do inter-zone transfer arrangements provide. No openly available tool addresses these questions at the SMA level, the geographic unit at which Japanese medical plans are written.

**Methods:** We developed MeshScope-Scenario, a probabilistic capacity–demand framework operating on the MeshScope-Region platform. For a selected SMA, observed inputs (notified ICU/HCU beds from the Hospital Bed Function Reports; resident population) are combined with four explicitly flagged assumption parameters — effective staffed-bed rate, concurrent severe-care demand per 100,000 population, surge multiplier, and net cross-boundary inflow — each with a user-specified distribution. A seeded Monte Carlo engine (deterministic reproduction under a fixed seed) estimates the distribution of bed shortfall; interventions are compared under common random numbers. Parameter dependence is introduced by a Gaussian copula with automatic positive-semidefinite correction; global sensitivity is quantified by Sobol first-order and total-order indices (Saltelli sampling, Jansen estimators) alongside a deterministic one-at-a-time tornado analysis. A two-zone extension transfers unmet demand to the nearest ICU-holding SMA using road-network travel times measured in MeshScope-Region, with a transfer time limit and an acceptance cap; because both zones share the same systemic draws, correlated exhaustion of donor capacity under surge (“shared-fate” risk) is represented structurally. A seasonal layer applies twelve monthly surge multipliers and reports the distribution of annual maximum shortfall and month-specific shortfall probabilities. The engine is a dependency-free pure-function module verified by 40 statistical tests.

**Results:** The framework reproduces identical output under identical seed and input; a flat seasonal profile reproduces the non-seasonal model exactly; copula factorization error is below 10^−9^; and 10,000 iterations across three intervention variants complete in approximately 50 ms in a standard browser, permitting fully interactive use. Applied to three archetypal SMAs from the observed FY2024 supply map (seed 42, 20,000 iterations, demand prior 5 per 100,000), an ICU-zero zone with a transfer partner 48 minutes away has shortfall probability 66.0% (P50 4.8, P90 20.7 beds); a median metropolitan zone, 32.7% (P90 5.9), with Sobol indices ranking demand density and surge dominant; a high-supply zone shows zero shortfall up to approximately 2.5× surge. For the ICU-zero zone, independent-donor reasoning credits the transfer arrangement with a 2.27-bed reduction in expected shortfall, of which shared-fate correlation removes 93%; the probability of severe shortfall under the arrangement equals that with no arrangement at all, while a committed pool of five donor beds (6% of donor effective supply) restores a 13-point reduction and more than halves the arrangement’s correlation exposure.

**Conclusions:** MeshScope-Scenario extends SMA-level capacity mapping from description to prospective risk assessment. All demand-side inputs are declared assumptions with adjustable distributions rather than estimates presented as fact; the framework’s value is to make the consequences of those assumptions, and their interaction with observed supply, explicit, reproducible, and inspectable for planning deliberation.

## 1. Introduction

Japanese medical plans, including the regional healthcare vision (chiiki iryo koso), are written at the level of secondary medical areas (SMAs). Recent work has begun to characterize critical care supply at this level: Ohbe and colleagues documented the national distribution of SMAs without ICUs and the outcomes of their residents [2, 3], and our companion study, MeshScope-Region, mapped the full ICU and high care unit (HCU) supply structure of all SMAs, its road-network accessibility, and its nine-year evolution, publishing the result as a standing platform [1].

These descriptive layers answer where capacity exists. Planning deliberation, however, also turns on prospective questions that description cannot answer: What is the probability that a given SMA’s critical care capacity is exceeded under a demand surge? Which of the quantities we do not observe — effective staffing, background demand, surge intensity, patient flows — dominates that risk? And how much protection does an inter-zone transfer arrangement actually provide when a surge affects donor and recipient zones simultaneously? These are questions about uncertainty, and answering them honestly requires a probabilistic treatment in which unobserved quantities are declared as assumptions with distributions, not presented as point estimates.

Deterministic bed-requirement formulas of the kind used in plan preparation do not expose this uncertainty, and patient-level outcome studies [3] condition on the existing supply map rather than exploring counterfactual capacity states. Simulation modeling is the standard instrument for such questions, and good-practice guidance for its conduct and reporting is well established [8, 9]; what has been missing is an implementation at the SMA level, connected to observed Japanese supply data, and open enough to be audited.

We therefore developed MeshScope-Scenario, a seeded Monte Carlo capacity–demand framework built directly on the MeshScope-Region platform. This paper specifies the model, its dependence structure, its sensitivity-analysis machinery, its two-zone transfer and seasonal-surge extensions, and its verification; it then illustrates the framework on archetypal SMAs drawn from the observed fiscal-year-2024 supply map. The framework is released with source code and a browser-based interface so that any stated result can be reproduced from its seed.

## 2. Methods

### 2.1 Design overview

MeshScope-Scenario is a probabilistic module operating on MeshScope-Region [1]. Region supplies the observed layer: notified ICU and HCU beds by SMA from the Hospital Bed Function Reports (byosho kino hokoku), resident population, and road-network travel times between SMAs. Scenario supplies the stochastic layer: a capacity–demand model whose unobserved inputs are explicit assumption parameters with user-specified distributions. The separation is enforced in the interface: every assumption parameter carries a visible “assumed” badge, and observed inputs cannot be confused with priors.

### 2.2 Observed inputs

For a selected SMA, the engine receives the notified ICU (or ICU+HCU) bed count and resident population from the Region platform (Hospital Bed Function Reports, fiscal year 2024; Basic Resident Registration population). For the transfer extension it additionally receives, for the nearest ICU-holding SMA, that zone’s beds, population, and the road-network travel time in minutes computed in Region with the Open Source Routing Machine [1]. All observed inputs are public administrative data; no patient-level data are used anywhere in the framework.

### 2.3 Capacity–demand model

The core model is a single-zone steady-state approximation. Effective supply is the notified bed count multiplied by an effective staffed-bed rate, plus any intervention bed change. Effective demand is (population / 100,000) multiplied by a concurrent severe-care demand rate per 100,000, a surge multiplier, and (1 + net cross-boundary inflow rate), reduced by any intervention demand-reduction fraction. Shortfall is max(0, demand − supply); surplus is max(0, supply − demand). Four parameters are treated as uncertain, with the following default priors (all adjustable, and all flagged as assumptions in the interface): effective staffed-bed rate, triangular (min 0.70, mode 0.85, max 0.95), reflecting the absence of public staffing-adjusted occupancy data; concurrent severe-care demand, normal (mean 8, SD 2) per 100,000; surge multiplier, uniform (1.0, 1.5); and net inflow rate, normal (mean 0, SD 0.05), pending connection of observed patient-flow data. Distribution families available are fixed, normal, uniform, and triangular, with non-negativity clamps where applicable.

### 2.4 Monte Carlo estimation and common random numbers

The engine draws parameter vectors with a seeded pseudo-random generator (mulberry32); identical input and seed reproduce identical output to the last digit, on any machine. Between 100 and 100,000 iterations may be requested; 10,000 iterations with three intervention variants complete in approximately 50 ms in a standard browser. Interventions (bed additions, demand-reduction measures such as tele-ICU support or wide-area transport coordination) are evaluated under common random numbers: every variant is scored on the same draw, so that Monte Carlo noise cancels from between-variant comparisons. Outputs per variant comprise the shortfall probability, a severe-shortfall probability against a user-set bed threshold, the full quantile set (P5–P95), expected shortfall and surplus, a histogram, and an empirical distribution function; complete results are exportable as JSON including the seed and model version for audit.

### 2.5 Dependence structure (Gaussian copula)

Parameters may be correlated — for example, surge intensity and reduced effective staffing may co-occur. Dependence is introduced by a Gaussian copula: correlated standard normal draws are generated by Cholesky factorization of a user-edited 4 × 4 correlation matrix and transformed to each parameter’s marginal by inverse-CDF mapping, so marginal distributions are preserved exactly. If the entered matrix is not positive semidefinite, it is corrected to the nearest valid correlation matrix by eigenvalue clipping, and the correction is disclosed in the interface and in the exported results rather than applied silently.

### 2.6 Sensitivity analysis

Two complementary instruments are provided. A deterministic one-at-a-time tornado analysis evaluates the shortfall at each parameter’s P10 and P90 with all others held at their medians, giving an exact, sampling-free local ranking. Global sensitivity is quantified by variance-based Sobol indices: first-order (S1) and total-order (ST) indices are estimated from Saltelli A/B/AB sampling with Jansen estimators [4–6], at n = 2,048 base samples under a dedicated seed. Sobol estimation assumes parameter independence; when the copula is enabled, indices are therefore reported explicitly as “indices under an independence assumption,” and this caveat is displayed with the results. Sensitivity analyses are evaluated on the core single-zone model, without the transfer and seasonal layers, and are labeled accordingly.

### 2.7 Inter-zone transfer extension (two-zone approximation)

When enabled, unmet demand in the index SMA is transferred to the nearest ICU-holding SMA, subject to (i) a transfer feasibility condition on road travel time (default limit 120 minutes; travel times are Region’s road-network measurements, not straight-line approximations), and (ii) an acceptance cap in beds. The donor zone’s own supply and demand are evaluated on the same systemic parameter draw as the index zone — the same staffed-bed rate, demand density, and surge realization — so that the model structurally represents correlated exhaustion: precisely when the index zone is stressed, the donor’s surplus is thinner. Transferred volume per iteration is the minimum of the index shortfall, the donor surplus, and the cap; expected transfer volume is reported. We emphasize the deliberate scope restriction: this is a two-zone approximation for planning screening, not a network equilibrium over all SMAs, and the interface states this limitation.

### 2.8 Seasonal surge extension

When enabled, twelve monthly multipliers scale the surge draw (presets: flat, summer-peaked, winter-peaked; the vector is editable and provides the connection point for external environmental series such as WBGT-based heat-risk profiles). Within each iteration all twelve months share the annual systemic draw, and outputs are extended to the distribution of the annual maximum shortfall and the month-specific shortfall probabilities. With a flat profile the seasonal model reproduces the non-seasonal model exactly, which is enforced by test.

### 2.9 Verification and reproducibility

The engine is a dependency-free pure-function TypeScript module accompanied by 40 statistical verification tests, including: exact reproduction under identical seeds; distribution-sampling checks; copula factorization accuracy (reconstruction error of the correlation matrix below 10^−9^) and tail behavior under positive correlation; Sobol properties (zero indices for fixed parameters, seed reproducibility, dominance of demand-side parameters under default priors); transfer-model properties (expected shortfall decreases with a surplus donor, is unchanged when the cap is zero or the time limit is exceeded, non-negativity); and exact flat-profile equivalence of the seasonal layer. All analyses in this paper state their seed and iteration count and are reproducible from the released code.

### 2.10 Ethics

The framework uses exclusively public, aggregate administrative data (notified beds, population, road networks). No patient-level or facility-confidential data are used, and no ethics approval is required. Outputs are planning-support estimates conditional on declared assumptions and are not intended for, and must not be used in, individual clinical or dispatch decisions.

## 3. Results

### 3.1 Framework properties

Determinism, performance, and correctness properties are established by the verification suite (Section 2.9): identical seeds reproduce identical results; 10,000 iterations across three variants complete in approximately 50 ms, allowing assumption distributions to be adjusted interactively during deliberation; the copula preserves marginals with factorization error below 10^−9^; and the seasonal layer is exactly consistent with the core model under a flat profile. All 40 tests pass in the released version (scenario-poc-0.3).

### 3.2 Illustrative application to archetypal SMAs

We apply the released engine to three archetypal SMAs selected from the observed FY2024 supply map (notified beds and population from the national metrics table; road travel times computed on facility-point coordinates). All results use seed 42 and 20,000 iterations.

Because the default demand prior (8 per 100,000) places low-supply zones at the supply– demand break-even point, shortfall probabilities saturate and lose discriminating power there (for archetype (a): P(shortfall) = 95.5% under the default versus 66.0% under 5 per 100,000, and 36.8% under 4). We therefore report archetype results under a demand prior of Normal(5, 1.25) per 100,000 and treat the saturation itself as a finding: conclusions in low-supply zones are structurally sensitive to the demand prior, which is precisely why the tool exposes it as an editable assumption rather than a fixed constant. Table 1 summarizes inputs and headline outputs; Figure 1 shows the shortfall distributions.

**Table 1.** Archetype inputs and baseline outputs (seed 42, 20,000 iterations, demand prior Normal(5, 1.25) per 100,000, summer-peaked seasonal profile; archetype (a) shown without transfer).

| Archetype | Zone | Population | ICU | HCU | per 100k | P(shortfall) | P(severe) | E[shortfall] | P10/P50/P90 |
| --- | --- | --- | --- | --- | --- | --- | --- | --- | --- |
| (a) ICU-zero | Chuetsu (1504) | 410,249 | 0 | 40 | 9.8 | 66.0% | 31.4% | 7.29 | 0 / 4.8 / 20.7 |
| (b) Median | Matsue (3201) | 229,425 | 14 | 15 | 12.6 | 32.7% | 2.7% | 1.43 | 0 / 0 / 5.9 |
| (c) High-supply | Maebashi (1001) | 329,120 | 41 | 88 | 39.2 | 0.0% | 0.0% | 0.00 | 0 / 0 / 0 |

**Figure 1.**
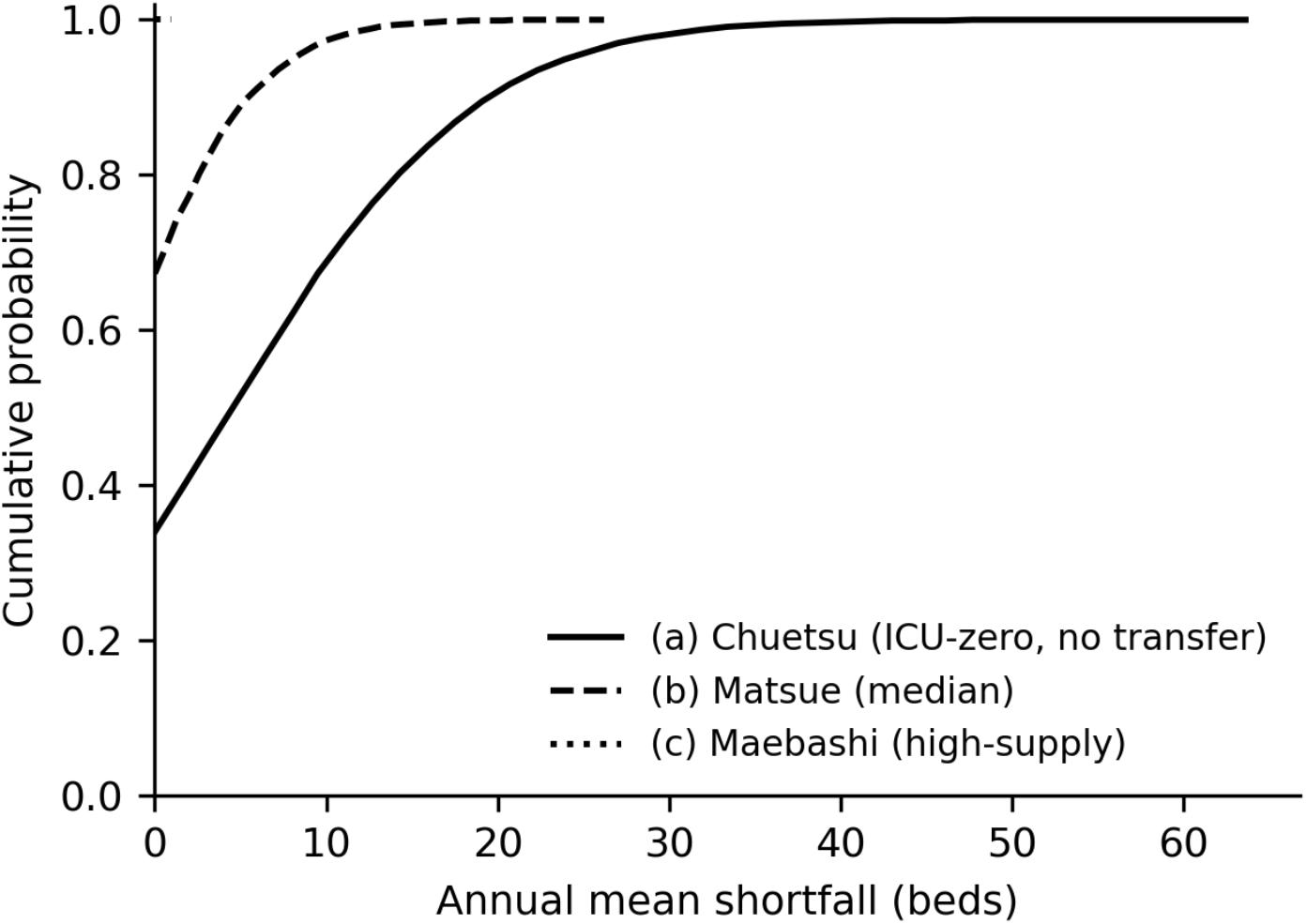
Cumulative distributions of annual mean shortfall for the three archetypes (archetype (a) without transfer).

**Figure 2.**
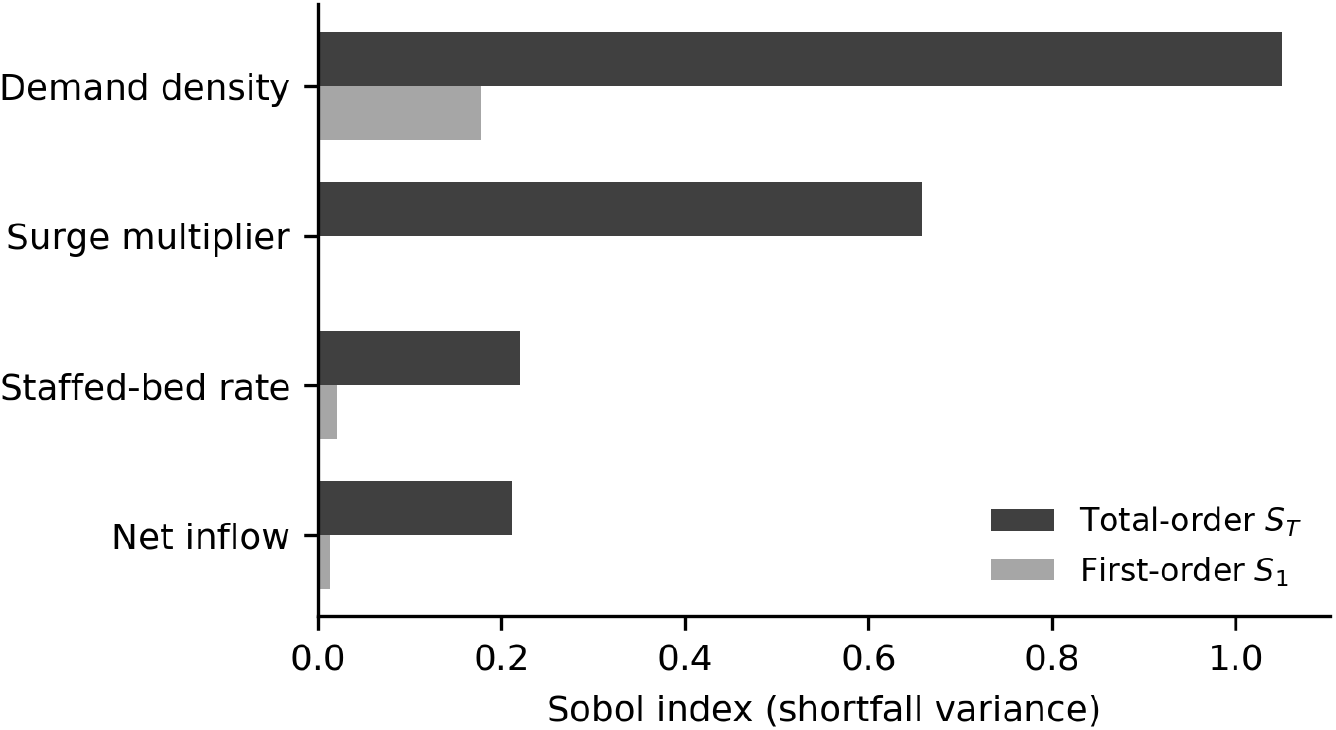
First-order and total-order Sobol indices for archetype (b) (Matsue). The one-at-a-time tornado is degenerate in this zone (zero shortfall at all OAT points), motivating variance-based indices.

Archetype (a): Chuetsu reports no ICU-classified beds and 40 HCU beds for 410,249 residents; its nearest ICU-holding partner, Niigata (18 ICU + 74 HCU), is 48 minutes away by road. Without transfer, annual expected shortfall is 7.29 beds (CVaR_95_ 29.69) and the probability of a two-month persistent severe episode is 26.9%. Under the summer-peaked profile, monthly shortfall probability ranges from 7.1% in winter to 66.0% at the August peak (Figure 3); enabling transfer under the shared-fate default shifts the August peak to 57.7% — the correlation-dependence of this modest improvement is quantified in Section 3.3.

**Figure 3.**
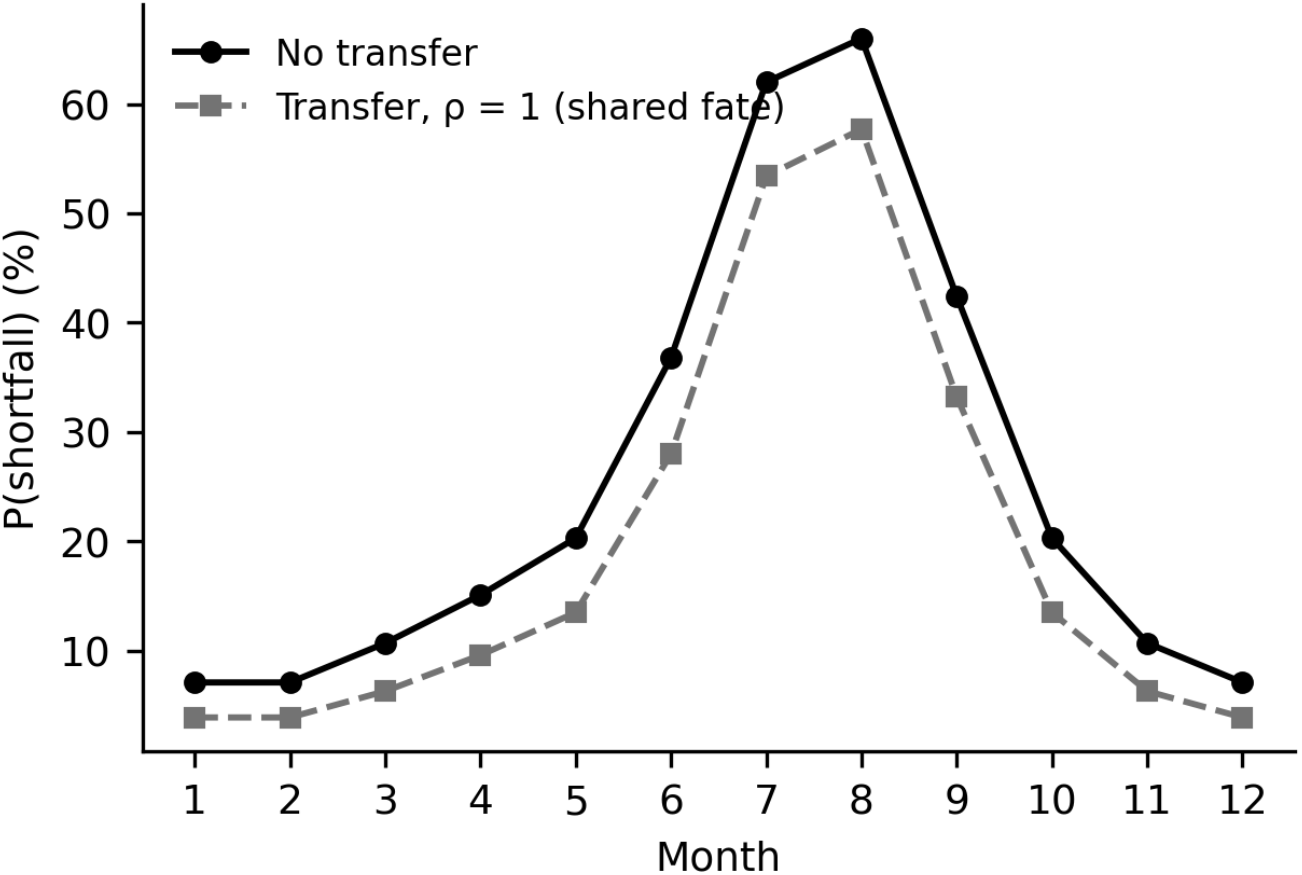
Monthly shortfall probability for archetype (a) under the summer-peaked profile, without transfer and with transfer under the shared-fate default (ρ = 1).

Archetype (b): Matsue sits at the national median of combined supply (12.6 per 100,000, exactly the national median). Baseline shortfall probability is 32.7% with expected shortfall 1.43 beds (P50 = 0, P90 = 5.9); severe shortfall is rare (2.7%). The one-at-a-time tornado is degenerate here — with all other parameters at their medians the zone has zero shortfall at every OAT evaluation point, so deterministic sensitivity bars vanish — which is itself the reason variance-based indices are required. Sobol indices on the stochastic output (Figure 2) rank demand density dominant (S_T = 1.05; total-order estimates can slightly exceed 1 under estimator variance), surge second (S_T = 0.66), with staffed-bed rate (0.22) and net inflow (0.21) minor, matching the expected ordering.

Archetype (c): Maebashi (39.2 per 100,000) shows zero shortfall probability under default priors. Sweeping a narrow surge band across multipliers, risk remains at zero through 2.0×, becomes measurable at 2.5× (0.4%), and material only around 2.75–3.0× (1.8% and 4.7%): the high-supply archetype absorbs roughly two and a half times the reference surge before shortfall risk appears.

### 3.3 Effect of the transfer arrangement under correlated surge

For archetype (a) we compare no transfer, transfer with independent draws (ρ = 0; the counterfactual implicit in independent-donor reasoning), and transfer with common systemic draws (ρ = 1; the model default), holding all randomness fixed across arms (seed 42, 20,000 iterations). Expected transfer volume erodes monotonically from 0.97 beds at ρ = 0 to 0.09 at ρ = 1 (−91%). The gap between the two transfer rows is the quantity of interest: independent-donor reasoning credits the arrangement with a 2.27-bed reduction in expected shortfall (7.29 → 5.02) and a 4.2-bed reduction in P90 shortfall (20.7 → 16.5), of which shared exposure removes 93% and 100% respectively (7.29 → 7.14; P90 20.7 → 20.7). For threshold measures the erosion is complete: severe-shortfall probability under the arrangement at ρ = 1 (31.4%) and the probability of a two-month persistent episode (26.9%) are indistinguishable from having no arrangement at all (Table 2; Figure 4A). All risk functionals worsen monotonically in ρ in this configuration; in deeply saturated zones the direction of threshold measures can reverse, so mass-based and threshold-based indicators should be read together.

**Table 2.**
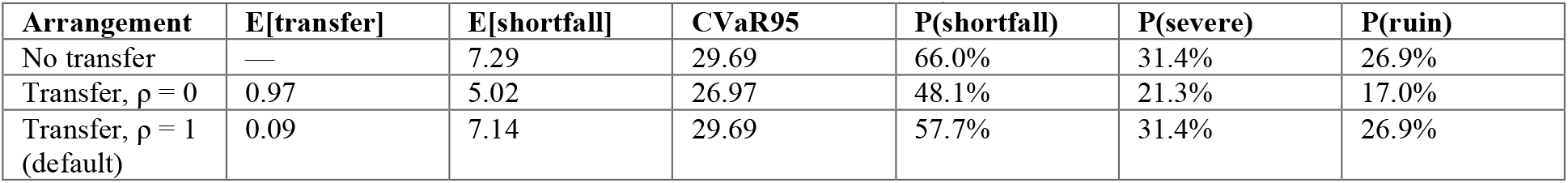
Transfer arrangement under inter-zone correlation, archetype (a) (seed 42, 20,000 iterations; transfer cap 20 beds; severe threshold 10 beds; ruin = severe shortfall in two consecutive months).

| Arrangement | E[transfer] | E[shortfall] | CVaR95 | P(shortfall) | P(severe) | P(ruin) |
| --- | --- | --- | --- | --- | --- | --- |
| No transfer | — | 7.29 | 29.69 | 66.0% | 31.4% | 26.9% |
| Transfer, $\rho = 0$ | 0.97 | 5.02 | 26.97 | 48.1% | 21.3% | 17.0% |
| Transfer, $\rho = 1$ (default) | 0.09 | 7.14 | 29.69 | 57.7% | 31.4% | 26.9% |

**Figure 4.**
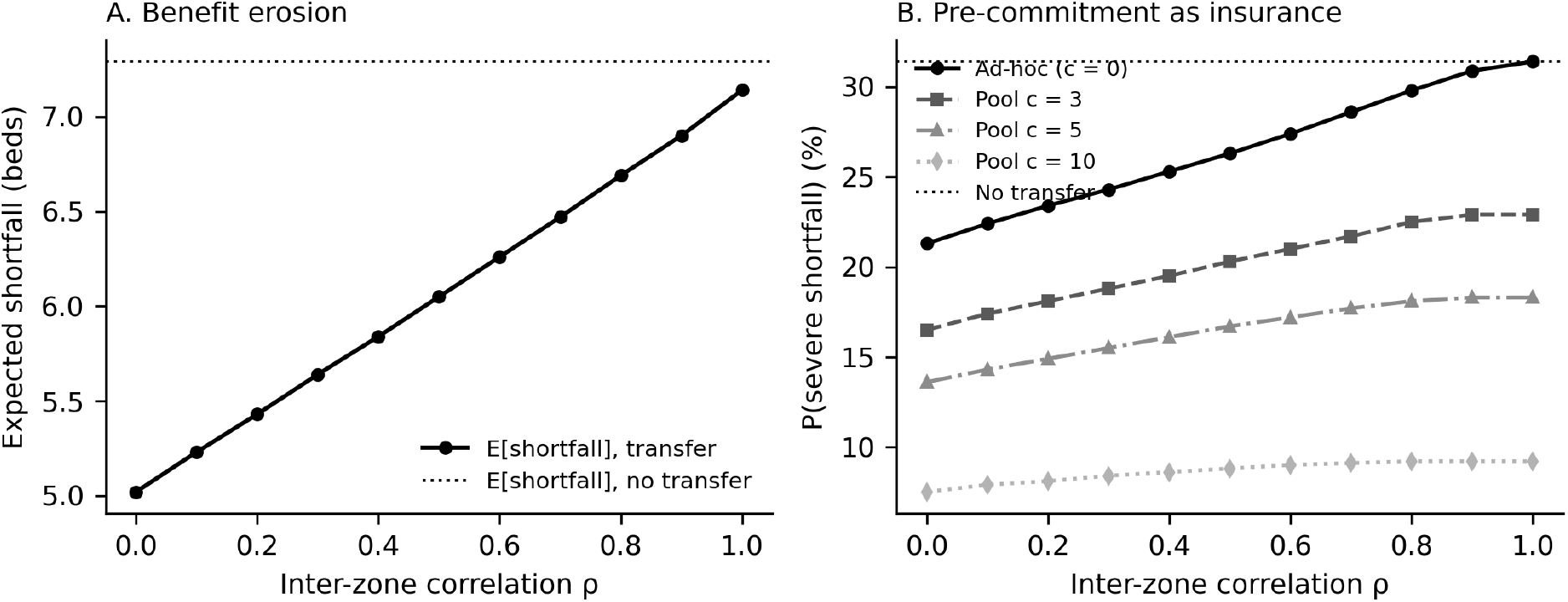
(A) Expected shortfall under the transfer arrangement as a function of inter-zone correlation ρ, against the no-transfer reference. (B) Severe-shortfall probability by arrangement type: ad-hoc transfer versus committed pools of c beds; pool curves flatten toward ρ-invariance.

The erosion is a property of the arrangement type, not of cooperation as such. A committed regional pool — c beds carved out of donor capacity in advance and available regardless of the donor’s own draws, the mechanism of Litvak et al.’s jointly reserved regional beds — is structurally invariant to ρ. Whereas the ad-hoc arrangement’s crisis-time value is measured at zero, a committed pool of five beds (6% of the donor’s effective supply) restores a 13-point reduction in severe-shortfall probability (31.4% → 18.3%) and cuts the arrangement’s correlation exposure by more than half (ΔP(severe) from ρ = 0 to 1: +10.1 points at c = 0, +4.7 at c = 5, +1.7 at c = 10; Figure 4B). Pre-commitment, in this precise sense, is insurance against correlation collapse, and its premium is a small, known fraction of donor capacity.

Most Japanese inter-zone arrangements are of the ad-hoc type.

## 4. Discussion

### 4.1 Principal contribution

MeshScope-Scenario converts an observed SMA-level supply map into an instrument for prospective risk assessment. Its design commitments are methodological rather than empirical: unobserved quantities are declared assumptions with distributions and visible flags; every stochastic result is reproducible from a stated seed; intervention comparisons are protected from Monte Carlo noise by common random numbers; dependence and its consequences are modeled rather than ignored; and the model’s central structural insight — that transfer arrangements provide less protection under surge than independent-donor reasoning suggests, because donor and recipient share systemic conditions — is represented in the simulation mechanics themselves.

### 4.2 Relation to prior work

The framework is complementary to both strands of existing SMA-level evidence. Patient-level outcome studies [2, 3] estimate the consequences of the supply map as it exists; MeshScope-Region [1] characterizes that map descriptively; MeshScope-Scenario asks what would happen under states of the world that have not (yet) occurred. Relative to deterministic bed-requirement formulas used in plan preparation, the framework’s contribution is not different arithmetic but the explicit propagation of uncertainty and the attribution of output variance to specific assumptions via Sobol indices — following established good practice for decision-analytic modeling [8, 9]. The seasonal layer’s WBGT connection point links the framework to environmental surge forecasting, and the transfer layer operationalizes, in prospective form, the access mechanism that Region’s drive-time analysis measures descriptively.

### 4.3 Intended use and misuse

The intended user is a planning body deliberating over an SMA’s critical care resilience: the framework supports asking which assumptions matter, how robust a conclusion is to plausible priors, and what a transfer arrangement is worth. It is a screening and deliberation instrument. It is not a forecasting system — outputs are conditional on declared assumptions, not predictions — and it is not an operational dispatch tool. Presenting conditional planning estimates as forecasts would be a misuse, and the interface’s assumption badges, model-version stamps, and exported seeds are designed to make such misuse difficult.

### 4.4 Limitations

First, the model is a single-zone steady-state approximation with a two-zone transfer extension; it does not represent full inter-zone network flows, within-zone heterogeneity, or queueing dynamics, and annual-maximum seasonal results share one systemic draw across months. Second, results depend on assumption priors, by design; the default priors are starting points for deliberation, not estimates, and demand-side priors in particular should be localized before any zone-specific conclusion is drawn. Third, notified beds overstate operationally staffed capacity to an unknown, zone-varying degree; the staffed-bed rate parameter expresses this uncertainty but does not resolve it. Fourth, sensitivity indices quantify statistical, not causal, attribution, and Sobol estimation assumes independence. Fifth, the transfer model abstracts from transport resources, clinical transferability of patients, and inter-organizational agreements. Extensions under design — full network flows over the Region origin–destination matrix, observed patient-flow priors for the inflow parameter, and WBGT-driven seasonal profiles — address several of these limits.

### 4.5 Conclusions

Probabilistic, assumption-explicit, reproducible scenario analysis at the SMA level is feasible with public data and lightweight open tooling, and it changes the questions a planning body can ask — from “how many beds are there” to “how likely is failure, what drives it, and what would an arrangement be worth.” MeshScope-Scenario provides that instrument as an open, verifiable companion to the descriptive supply map.

## Declarations

### Data availability

All inputs are public administrative data: Hospital Bed Function Reports (Ministry of Health, Labour and Welfare), Basic Resident Registration population (Ministry of Internal Affairs and Communications, e-Stat), and road-network data © OpenStreetMap contributors (ODbL), routed with OSRM. No patient-level data are used. Zone-level supply metrics, facility-point coordinates for the 1,045 notifying facilities, 330-SMA boundaries, and road-network access indicators are deposited at Zenodo (version DOI: 10.5281/zenodo.21781052; concept DOI: 10.5281/zenodo.21781051).

### Code availability

The simulation engine (pure-function TypeScript, dependency-free; model version scenario-poc-0.3), the downside-risk extension riskx v0.4.0 used in Sections 3.2–3.3 (CVaR / expected shortfall, ruin probability, inter-zone correlation with common-random-number sweep, committed regional pool), and the verification suites (40 engine tests; 49 extension tests) are archived at Zenodo (version DOI: 10.5281/zenodo.21781516; concept DOI resolving to the latest version: 10.5281/zenodo.21781515) and maintained at https://github.com/star-observer2027/meshscope-riskx.

### Funding

This work received no external funding.

### Competing interests

K.O. is the representative director of Jinen Co., Ltd., which develops the MeshScope platform, and serves as staff of NPO ICON. The framework described here uses public data only.

### Author contributions

K.O. conceived the framework, implemented the engine and platform, designed the verification suite, and wrote the manuscript; co-authors advised on clinical and health-security framing and revised the manuscript.

## Notes

### Author Declarations

The framework uses exclusively public, aggregate administrative data (notified beds, population, road networks). No patient-level or facility-confidential data are used, and no ethics approval is required.

